# Differential Effects of Pathological Beta Burst Dynamics Between Parkinson’s Disease Phenotypes Across Different Movements

**DOI:** 10.1101/2020.04.12.20063032

**Authors:** Raumin S. Neuville, Matthew N. Petrucci, Kevin B. Wilkins, Ross. W. Anderson, Shannon L. Hoffman, Jordan E. Parker, Anca Velisar, Helen M. Bronte-Stewart

**Affiliations:** Stanford University School of Medicine, Department of Neurology and Neurological Sciences, Stanford, CA, USA; Stanford University School of Medicine, Department of Neurosurgery, Stanford, CA, USA; University of California School of Medicine, Irvine, CA, USA; University of California Los Angeles, Los Angeles, CA, USA; Smith Kettlewell Research Institute, San Francisco, CA, USA

**Keywords:** beta oscillations, Parkinson’s disease, local field potentials, subthalamic nucleus, deep brain stimulation, burst duration, movement, motor control

## Abstract

**Background:** Resting state beta band (13 – 30 Hz) oscillations represent pathological neural activity in Parkinson’s disease (PD). It is unknown how the peak frequency or dynamics of beta oscillations may change among fine, limb and axial movements and different disease phenotypes. This will be critical for the development of personalized closed loop deep brain stimulation (DBS) algorithms during different activity states.

**Methods:** Subthalamic (STN) local field potentials (LFPs) were recorded from a sensing neurostimulator (Activa® PC+S, Medtronic PLC.,) in fourteen PD participants (six tremor-dominant, 8 akinetic-rigid) off medication/off STN DBS during thirty seconds of repetitive alternating finger tapping, wrist-flexion extension, stepping in place, and free walking. Beta power peaks and beta burst dynamics were identified by custom algorithms and were compared among movement tasks and between tremor-dominant and akinetic-rigid groups.

**Results:** Beta power peaks were evident during fine, limb, and axial movements in 98% of movement trials; the peak frequencies were similar during each type of movement. Burst power and duration were significantly larger in the high beta band, but not in the low beta band, in the akinetic-rigid group compared to the tremor-dominant group.

**Conclusions:** The conservation of beta peak frequency during different activity states supports the feasibility of patient-specific closed loop DBS algorithms driven by the dynamics of the same beta band during different activities. Akinetic-rigid participants had greater power and longer burst durations in the high beta band than tremor-dominant participants during movement, which may relate to the difference in underlying pathophysiology between phenotypes.

## Introduction

Exaggerated resting state beta band (13 – 30 Hz) oscillations and synchrony are pathophysiological markers of hypokinetic aspects of Parkinson’s disease (PD). When averaged over time, these oscillations appear as elevated portions of the local field potential (LFP) power spectral density (PSD) above the broadband 1/f spectrum (He, 2014; Shreve et al., 2017). Beta band power is attenuated on dopaminergic medication and during subthalamic (STN) deep brain stimulation (DBS); the degree of attenuation has been correlated to the degree of improvement in bradykinesia and rigidity, whereas averaged resting state beta band power is less robustly correlated with PD motor signs (Bronte-Stewart et al., 2009; Brown et al., 2001; Cassidy et al., 2002; Eusebio et al., 2011; Kehnemouyi et al., 2021; Kühn et al., 2009, 2008, 2006; Levy et al., 2002; Priori et al., 2004; Quinn et al., 2015; Ray et al., 2008; Weinberger et al., 2006; Whitmer et al., 2012; Williams et al., 2002).

Recently, it has been shown that physiological resting state beta oscillations are represented by short duration fluctuations in power (beta bursts) in the striatum and cortex of healthy non-human primates (Feingold et al., 2015). These authors suggested that the precise temporal dynamics of beta bursts may be more reliable markers of PD than averaging beta activity over periods of time. Burst dynamics in PD have been studied during rest (RW Anderson et al., 2020; Tinkhauser et al., 2017), but less is known about real time beta burst dynamics during movement and whether beta burst dynamics differ during fine motor or limb movements and/or during gait and freezing of gait (FOG) (Anidi et al., 2018; Lofredi et al., 2019, Kehnemouyi et al., 2020). The duration of beta bursts is a relevant neural control variable for closed loop DBS systems, which can precisely target (shorten) the duration of beta bursts, but it is not known how this variable may change among movements which may necessitate a different response from a closed-loop algorithm (Petrucci et al., 2020a).

In addition to differences among tasks, it is unclear how beta burst dynamics may differ between sub bands of beta or between Parkinson’s disease phenotypes. Previous studies that have evaluated phenotype differences primarily focused on high and low beta band power in the operating room or perioperative state (i.e., the week after implantation). Differences in high beta band power were demonstrated between tremor-dominant and akinetic-rigid subtypes at rest, but not during movement in an elbow-flexion task in the operating room (Godinho et al., 2021). Furthermore, within band differences between rest and movement were observed for each subtype (low beta for tremor dominant, high beta for akinetic rigid). Differences in resting state high beta power have also been reported in the immediate post-operative period between people with and without freezing of gait, as assessed off medication in the pre-operative period (Toledo et al., 2014). To date, no study has compared burst durations within sub bands of beta, between disease phenotypes, and during different movements using a chronically implanted device. In this study, we investigated whether beta band peak frequencies were conserved or were different during fine, limb, and/or axial movements in people with PD, and whether there were differences in beta band and sub band power and burst dynamics between the akinetic-rigid and tremor-dominant phenotypes.

## Methods

### Human Participants

Fourteen participants (10 male) with clinically established Parkinson’s disease (PD) underwent bilateral implantation of DBS leads (model 3389, Medtronic, PLC, Minneapolis, MN, USA) in the sensorimotor region of the subthalamic nucleus (STN) using a standard functional frameless stereotactic technique and microelectrode recording (MER) (Brontë-Stewart et al., 2010; Quinn et al., 2015). Long-acting dopaminergic medication was withdrawn over 24 hours (72 hours for extended-release dopamine agonists) and short-acting medication was withdrawn for over 12 hours before surgery and before each study visit. One participant took an extra short-acting carbidopa/levodopa tablet 5 hours before the experiments and was included as their resting state LFP spectra were similar 6.25 hours and 8.5 hours later, suggesting resolution of an attenuating effect of medication on beta power (Trager et al., 2016). The preoperative selection criteria and assessment of participants have been previously described (Bronte-Stewart et al., 2009; de Solages et al., 2010; Taylor Tavares et al., 2005). The dorsal and ventral borders of each STN were determined using MER, and the base of electrode 0 of the Medtronic 3389 lead was placed at the MER defined ventral border of the STN (de Solages et al., 2011, 2010; Marceglia et al., 2006). The DBS leads were located in the STN (Figure 2A). All participants signed a written consent and the study was approved by the Food and Drug Administration (FDA), Investigational Device Exemption (IDE) and the Stanford School of Medicine Institutional Review Board (IRB). Each participant was classified as tremor dominant (TD) or akinetic rigid (AR) phenotype based on previously described criteria (Quinn et al., 2015; Shreve et al., 2017; Trager et al., 2016) and the more and less affected sides were determined by unilateral Unified Parkinson’s Disease Rating Scale (UPDRS) part III sub-scores.

### Experimental Protocol

All experiments were performed within two months after DBS lead placement in the off medication/off DBS state. Recordings were collected in the Stanford Human Motor Control and Neuromodulation Laboratory. Experiments started with a resting state recording, during which each participant sat still for 30 seconds. Participants completed four different movement tasks (Figure 1): (1) quantitative digitography (QDG) on an engineered keyboard (Bronte-Stewart et al., 2000; Taylor Tavares et al., 2005; Trager et al., 2015) (2) instrumented repetitive wrist-flexion extension (WFE) using wearable sensors (Koop et al., 2008, 2006; Louie et al., 2009), (3) stepping in place (SIP) on dual force plates (Nantel et al., 2011), and (4) free walking (FW). During the QDG task, participants were seated with their elbow flexed at approximately 90 degrees and the wrist was supported by a pad alongside a customized engineered keyboard. Visual and auditory feedback was minimized, as the participants had their eyes closed and wore headphones that played white noise to limit auditory feedback from the key tapping. With the index and middle fingers placed on individual keys, participants were instructed to tap each key in an alternating pattern as fast and regularly as possible for 30 seconds. For the instrumented rWFE task, participants were seated with their elbow flexed at approximately 90 degrees and the hand in the mid-pronated-supinated position before they were asked to flex and extend their wrists as fast as possible for 30 seconds. During the SIP task, participants were instructed to perform alternating stepping on dual force plates for 100 seconds. For the FW task, all participants walked for approximately one minute that included portions of forward walking and turns. All movements were self-paced.

**Figure 1.** The **(A)** quantitative digitography (QDG), **(B)** instrumented repetitive wrist-flexion extension (WFE), **(C)** stepping in place (SIP) and **(D)** forward walking (FW) tasks. ***<u>Please contact the authors if you are interested in this figure</u>***.

### Data acquisition and analysis

Local field potentials (LFPs) from the STN were recorded from the electrode pair of the DBS lead that had the greatest resting state beta band peak power and the least artifact (electrode pairs 0-2 or 1-3 of the Medtronic 3389 lead; Supplementary Information, Table S1). The pre-amplified LFP was high-pass filtered at 2.5 Hz and low-pass filtered at 100 Hz. LFP data was sampled at a rate of 422 Hz (10-bit resolution). The gains used for the experiments were set at 2,000 with a center frequency of 2.5 Hz. The uncompressed LFP data were extracted via telemetry using the Activa™ PC+S tablet programmer and then transferred to a computer for offline analysis in MATLAB (version 9.5, The MathWorks Inc. Natick, MA, USA). LFP data used for analysis was from the first 30 seconds of movement or from the maximum length of continuous movement without cueing. The power spectral density (PSD) diagrams were calculated using Welch’s method, which used a 1-second Hanning window with 50% overlap (Welch, 1967). The peak frequency in the beta band was detected using a peak detection algorithm (de Solages et al., 2010); if more than one peak was detected, the peak with the greatest power was chosen. In two movement episodes, the algorithm failed to detect a peak, which was evident on visual inspection.

### LFP burst dynamics determination

The method of determining the burst dynamics was adopted from Anderson et al., which uses a baseline threshold calculated from a portion of the PD LFP spectrum that corresponds to the power and burst dynamics of a simulated, physiological 1/f spectrum (RW Anderson et al., 2020). The baseline method captures a broader range of beta burst durations than high power burst detection methods. The band of interest was first filtered into equal consecutive 6 Hz overlapping bands, using a 6-Hz bandwidth, zero-phase 8th order Butterworth filter, and then squared. An envelope was formed by interpolating the consecutive peaks of the squared signal. The threshold for characterizing individual represented the baseline power calculated by averaging the median trough amplitudes from 5 consecutive overlapping 6 Hz bands in the 45-63 Hz PD gamma spectrum. In contrast to the elevated, beta frequency band of the PD spectrum, the higher frequency band is not elevated above the physiological LFP activity or 1/f signal, and contains burst dynamics resembled that of physiological neural activity (RW Anderson et al., 2020; He, 2014). The duration of a burst was calculated as the time between consecutive crossings of the envelope across the baseline threshold. The average power of each burst was also calculated (mean burst power).

### Statistics

The primary outcome variables were movement peak frequency, PSD power, mean burst power, and mean burst duration. PSD power, mean burst power, and mean burst duration were calculated separately for low beta (14-20 Hz) and high beta (22-28 Hz) frequency bands. We used 6 Hz bands to allow for equal comparison of burst durations between bands (RW Anderson et al., 2020). Normalization of all power values was completed through division by the average power of the squared and rectified signal in the 45-63 Hz frequency band during the resting state (Anidi et al., 2018). Independent t-tests were used to compare age, disease duration, and pre-operative UPDRS scores between the TD and AR phenotypes. One-way repeated measures ANOVAs compared peak frequencies in the PSDs and variation in power and burst metrics among the different movement tasks in high and low beta with each STN treated individually and PD phenotype as a between-subjects factor. Analyses were corrected for multiple comparisons using Bonferroni correction. In the presence of a violation of Mauchly’s test of sphericity, the Greenhouse-Geisser correction was applied. There was one trial per movement task for each participant.

## Results

Of the fourteen participants, six were classified as TD and eight were classified as AR. The age of the group (mean ± SD) was 57.0 ± 10.2 years (TD 60.4 ± 10.8 years; AR 54.4 ± 9.5 years), and the disease duration from symptom onset was 7.7 ± 3.7 years (TD 8.7 ± 4.3 years; AR 7.0 ± 3.2 years). UPDRS III scores (mean ± SD) in the pre-operative off- and on-medication state were 39.2 ± 14.8 (TD 44.7 ± 12.3; AR 37.6 ± 16.1) and 23.2 ± 14.1 (TD 19.2 ± 7.7; AR 25.4 ± 16.5), respectively. There were no significant differences in age, disease duration, or pre-operative UPDRS scores between participants classified as TD and AR (p > 0.05). The DBS leads were well placed within the STN, Figure 2A.

**Figure 2.**
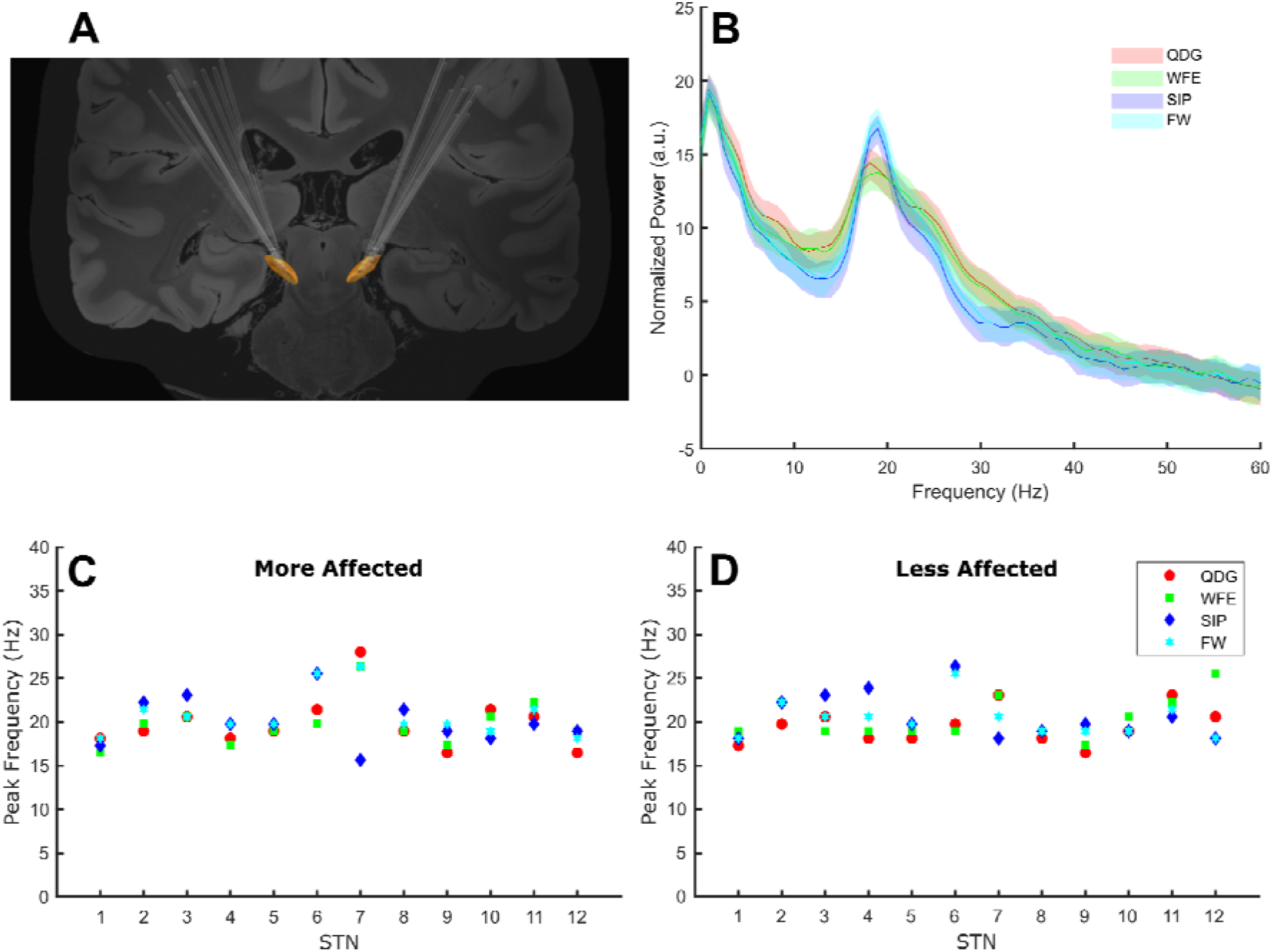
**(A)** The lead placements for all participants for the left and right STNs. **(B)** The normalized grand average power spectral density plots for the four tasks. **(C and D** The beta frequency peaks for each task by participant in the more and less affected STNs.

### Peak frequency was conserved across different movements

Among the cohort of 24 STNs (8 TD, 16 AR) for whom peaks could be detected, peaks of elevated beta power were detected in 98% of movement episodes during the different tasks, demonstrating that exaggerated beta band oscillations and synchrony were present during fine motor, limb and axial movements. In two TD participants, no peak was detected in either hemisphere, so they were excluded from this analysis. Beta peaks across the four movement tasks is depicted in the grand average PSDs in Figure 2B. The peak frequency did not differ across the movement tasks (F(1.62,35.53) = 0.58, p = 0.53, partial η^2^ = 0.026) or between phenotypes (F(1,22) = 0.39, p = 0.54, partial η^2^= 0.017), and there was no interaction between task and phenotype (F(1.62,35.53) = 2.93, p = 0.076, partial η^2^ = 0.12) on peak frequency. Peak frequency across movements was similar in the more (Figure 2C) and less affected (Figure 2D) STNs.

### Differences in PSD power between TD and AR groups in the high beta band

Normalized PSD power was analyzed across movement tasks and between the TD and AR groups in high and low beta for the full cohort of 28 STNs (Figure 3). In high beta, there was a significant effect of phenotype (F(1,26) = 8.84, p = 0.006, partial η^2^ = 0.25), but not of task (F(1.33,34.62) = 2.93, p = 0.085, partial η^2^ = 0.10) (Figure 3). Normalized high beta PSD power was greater for the AR group compared to the TD group across all movements (Figure 4A). In low beta, there were no differences in normalized PSD power between phenotypes (F(1,26) = 1.27, p = 0.270, partial η^2^ = 0.047) or across tasks (F(1.39,36.18) = 0.88, p = 0.39, partial η^2^ = 0.033). There was also no interaction between task and PD phenotype for normalized PSD power in either low beta (F(1.39,36.18) = 0.088, p = 0.85, partial η^2^ = 0.003) or high beta (F(1.33,34.62) = 0.59, p = 0.49, partial η^2^ = 0.022).

**Figure 3.**
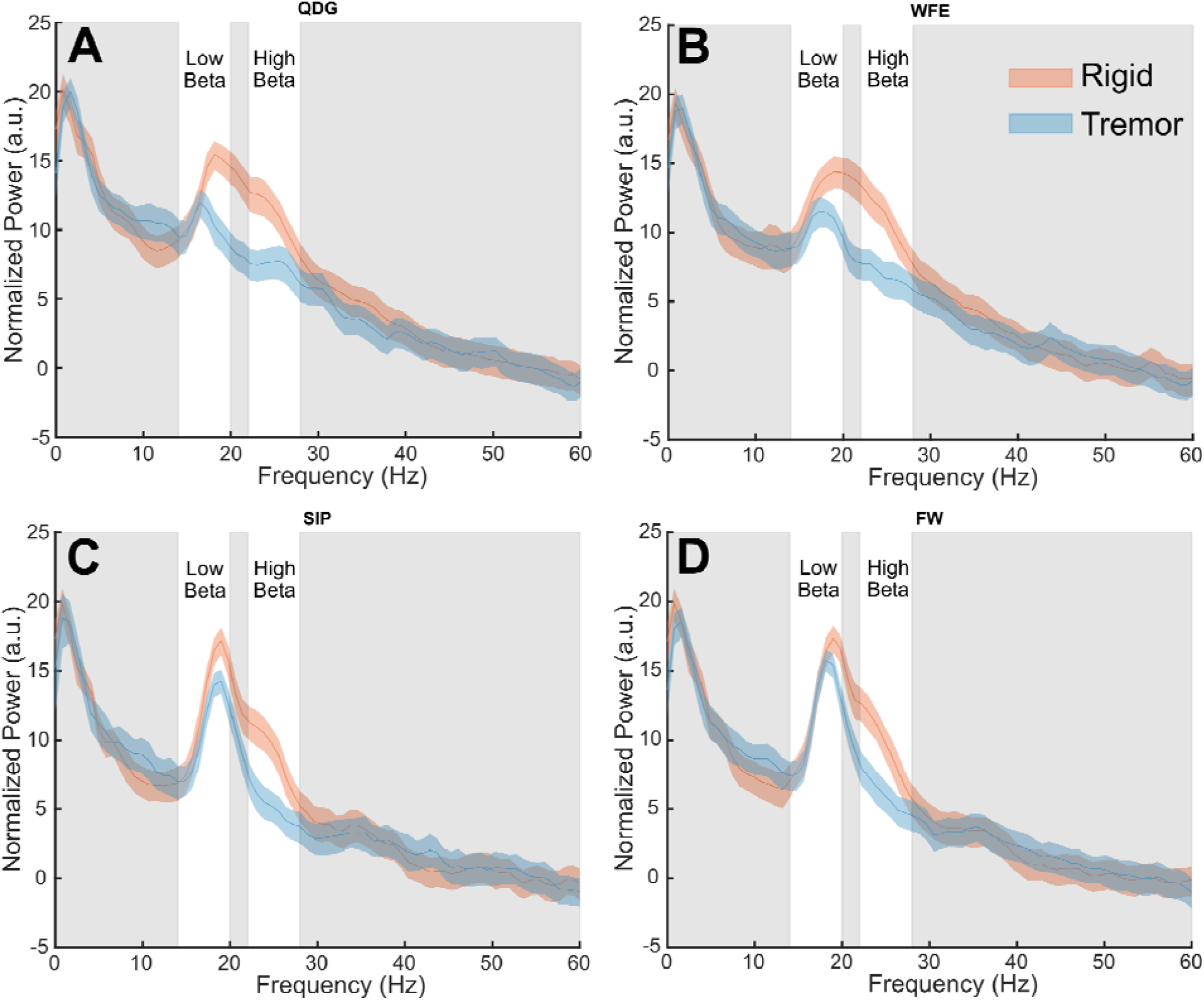
Grand average normalized power spectral density plots for each phenotype in the **(A)** QDG, **(B)** WFE, **(C)** SIP, and **(D)** FW tasks. There were significant differences (*p* < 0.05) between phenotypes in the high beta band.

**Figure 4.**
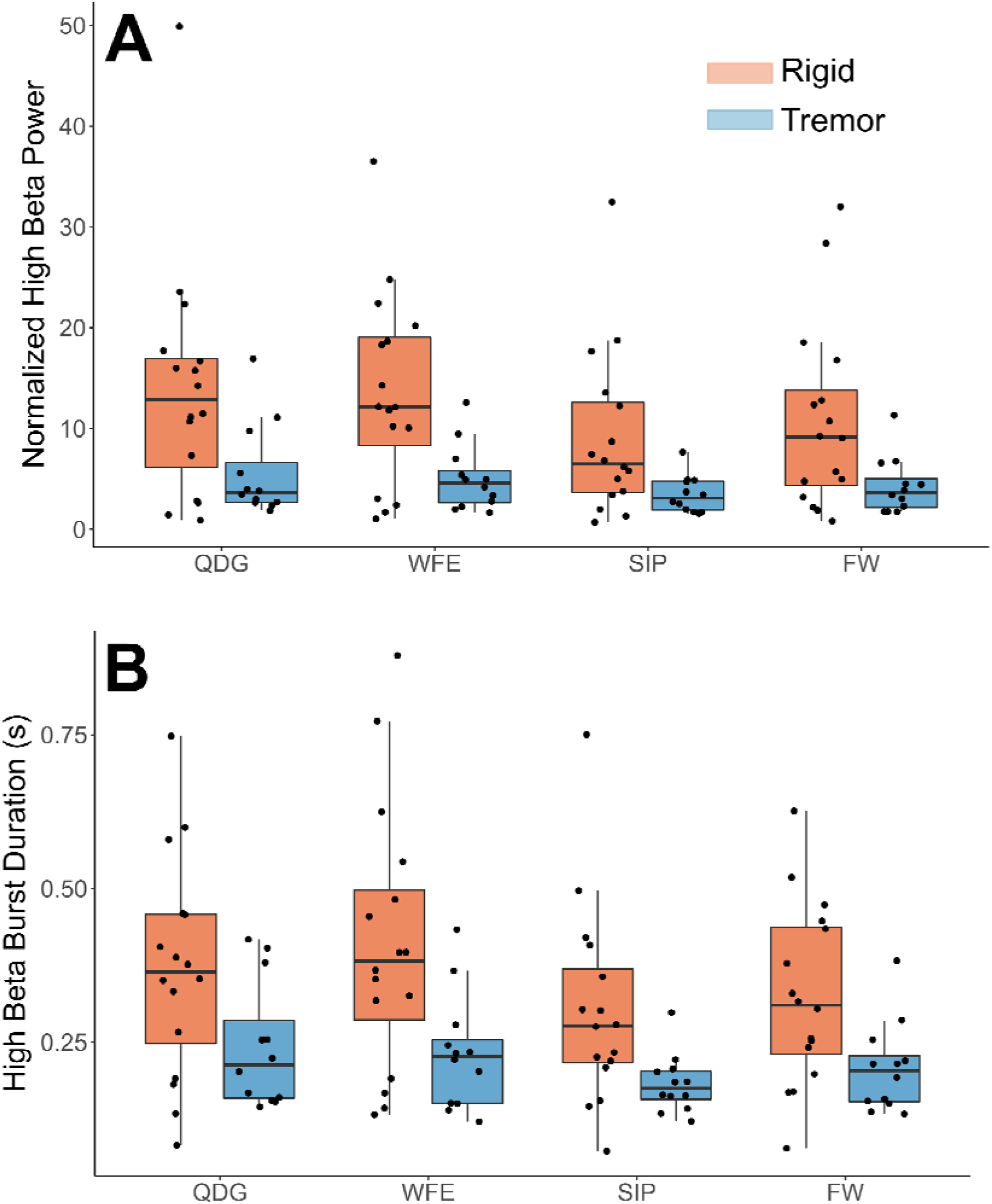
**(A)** Normalized high beta peak power and **(B)** high beta burst durations by task and phenotype. Note, a significant main effect was observed between groups (*p* < 0.05) but not task, and there was no significant interaction effect.

### Differences between the AR and TD groups and across tasks in high beta band

Mean burst duration was analyzed across movement tasks and between the TD and AR groups in high and low beta for 27 STNs (11 TD, 16 AR) (Figure 4B). Burst data for one STN of a TD patient was excluded because burst duration in low beta during the FW task was identified as a statistical outlier (greater than 3 SD from the mean). In high beta, there was both a significant effect of phenotype (F(1,25) = 8.92, p = 0.006, partial η^2^ = 0.26) and of task (F(1.75,43.67) = 4.48, p = 0.021, partial η^2^ = 0.15). High beta mean burst duration was greater for the AR phenotype compared to the TD phenotype across all movements (Figure 4B). Pairwise comparisons between movement tasks did not reveal significant differences between specific tasks with the Bonferroni correction (p > 0.05). In low beta, there were no differences in mean burst duration between phenotypes (F(1,25) = 2.34, p = 0.14), partial η^2^ = 0.085) or across tasks (F(1.36,33.87) = 0.18, p = 0.75, partial η^2^ = 0.007). There was no interaction between task and PD phenotype for mean burst duration for either low beta (F(1.36,33.87) = 0.44, p = 0.57, partial η^2^ = 0.017) or high beta (F(1.75,43.67) = 0.43, p = 0.63, partial η^2^ = 0.017).

### Differences in mean burst power between PD phenotypes, but not across tasks, in high beta

Mean burst power was analyzed across movement tasks and between PD phenotypes in high and low beta (Figure 4A). In high beta, there was a significant effect of phenotype (F(1,25) = 9.06, p = 0.006, partial η^2^ = 0.266), but no effect of task (F(1.34,33.50) = 0.41, p = 0.59, partial η^2^ = 0.016). High beta mean burst power was greater for the AR phenotype compared to the TD phenotype across all movements. In low beta, there were no differences in mean burst power between phenotypes (F(1,25) = 3.12, p = 0.090, partial η^2^ = 0.11) or across tasks (F(1.30,32.52) = 0.24, p = 0.69, partial η^2^ = 0.010). There was no interaction between task and PD phenotype for mean burst power in both low (F(1.30,32.52) = 0.084, p = 0.84, partial η^2^ = 0.003) and high (F(1.34,33.50) = 0.040, p = 0.90, partial η^2^ = 0.002) beta.

## Discussion

The results of this study demonstrate that pathological beta oscillations and synchrony are present during ongoing movement and that the frequencies of the beta band peak were similar among fine, limb and axial movements. However, people with PD classified as akinetic-rigid showed greater high beta power and high beta burst duration and burst power across all tasks compared to those classified as tremor-dominant. This difference may point to an important difference in pathophysiology between phenotypes.

### The clinical significance of the conservation of beta band peak frequency across movements

Several studies have demonstrated that beta power decreased before, at the onset of, and during movement in human participants with PD and in non-human primates (Anidi et al., 2018; Blumenfeld et al., 2017; Hell et al., 2018; Johnson et al., 2016; Joundi et al., 2013; Kühn et al., 2004; Litvak et al., 2011; Syrkin-Nikolau et al., 2017, Fischer 2018, Lofredi 2019). This has led to a frequent generalization in the literature that beta power ‘goes away’ during movement. The results of this study demonstrate that beta peaks were still evident during movement, and that the peak frequencies were conserved among fine motor and limb movements and during gait. This may alleviate concerns regarding the implementation of closed loop DBS in freely moving people. Up to now, closed loop DBS classifier algorithms have used estimates of resting state beta band power as the control variable (Afzal et al., 2019; Little et al., 2016a, 2016b, 2013; Petrucci et al., 2020b; Piña-Fuentes et al., 2019; Piña-Fuentes et al., 2017; Rosa et al., 2017, 2015; Velisar et al., 2019). Such algorithms require knowledge of the peak frequency of the band of interest and until now it was not known whether the same beta band could be used to drive closed loop DBS when the person is working at their computer, eating, dressing, or when walking. Although others have seen that there was a slight shift in peak frequency between different motor states (Canessa et al., 2020), we observed no significant difference across four different tasks. The conservation of the choice of the band of interest (determined by the peak frequency) among fine, limb, and axial movements suggests that the same classifier algorithms will be appropriate across movement states. Additionally, even if small differences are observed in peak frequencies, most current methods for tracking beta band look across a bandwidth of 6+/- Hz and therefore are robust against shifts in peak frequencies that still fall within these bandwidths (Afzal et al., 2019; Petrucci et al., 2020a; Velisar et al., 2019).

### Differences in pathophysiology between motor phenotypes

Our results demonstrate that the AR group shows greater high beta power and burst metrics across tasks compared to the TD group. This is the first study to show neural oscillatory differences in the STN between PD phenotypes across different movement states. High beta oscillations in the STN have been posited to relate to STN-cortical connections in PD, whereas low beta oscillations relate to intrinsic pathophysiology within the basal ganglia (Oswal et al., 2020). Specifically, coupling in high beta between the STN and supplementary motor area (SMA) correlates with fiber density between those two regions. Furthermore, improvement in rigidity with DBS has been shown to be related to connectivity to the SMA (Akram et al., 2017). The differences observed in our study between AR and TD may reflect differences in these STN-cortical interactions. This is further supported by the previous work demonstrating greater high beta power in freezers compared to non-freezers (Toledo et al., 2014) and tremor-dominant vs. akinetic-rigid (Godinho et al., 2021) at rest. Together, these results point to a pathological low beta oscillation that is consistent across phenotypes and then a potentially separate high beta oscillation that may be more specific to akinetic-rigid symptoms regardless of task. These differences could be utilized to improve patient-specific closed-loop loop algorithms due to recent advances in technology (Summit™ RC+S, Medtronic PLC) that can track multiple bands simultaneously.

## Limitations

Due to the limited number of investigative devices (Activa™ PC+S, Medtronic PLC, MN, USA) allocated to centers, the sample size was small but comparable to previous studies (Anidi et al., 2018; Blumenfeld et al., 2017; Quinn et al., 2015; Syrkin-Nikolau et al., 2017). Additionally, the tremor-dominant cohort displayed a mix of presence versus absence of tremor across the tasks and therefore it is difficult to say with certainty that the observed differences in high beta are a phenological difference between phenotypes or that the action of the tremor itself is specific to high beta. We did confirm in at least 2 participants that there was not an appreciable difference in high beta during the tremor and non-tremor periods when tremor arose in the middle of the trial (See Supplementary Information, Fig. S1-8). A larger cohort of tremor-dominant participants is needed to confirm these findings.

## Conclusion

The results of this study demonstrated that exaggerated beta power was evident during fine motor, limb and axial movements and that the peaks of the frequency band of elevated power were similar during such different movements. Furthermore, there were significant differences in beta power and burst durations between the akinetic-rigid and tremor-dominant phenotypes in the high beta, but not low beta. These findings are critical for future closed loop DBS systems, which will require an input that is both indicative of the disease state as well as robust through the patient’s activities of daily living.

## Data Availability

The datasets analysed during the current study are available from the corresponding author on reasonable request.

## Acknowledgements

We would like to thank Johanna O’Day, Muhammad Furqan Afzal, Thomas Prieto and the rest of the members of the Human Motor Control and Neuromodulation lab, and most importantly, the participants who dedicated their time to this study.

## Author Contributions

**Raumin Neuville:** Conceptualization, Methodology, Software, Validation, Formal analysis, Investigation, Data Curation, Writing – Original Draft and Review & Editing, Visualization, Supervision, Project administration. **Matthew Petrucci:** Conceptualization, Methodology, Validation, Formal analysis, Writing – Review & Editing, Supervision. **Kevin Wilkins:** Formal Analysis, Writing – Review & Editing, Visualization. **Ross Anderson:** Methodology, Software, Validation, Formal analysis, Investigation, Writing – Original Draft and Review & Editing, Visualization, Supervision. **Shannon Hoffman:** Formal Analysis, Writing-Review & Editing, Visualization. **Jordan Parker:** Methodology, Validation. **Anca Velisar:** Software, Investigation. **Helen Bronte-Stewart:** Conceptualization, Methodology, Writing – Original Draft & Review & Editing, Supervision, Funding acquisition.

