## Supplementary Information, Table S1 for "Differential Effects of Pathological Beta Burst Dynamics Between Parkinson’s Disease Phenotypes Across Different Movements"

| **Participant** | **LSTN** | **RSTN** |
| --- | --- | --- |
| 1 | 0-2 | 8-10 |
| 2 | 0-2 | 8-10 |
| 3 | 0-2 | 8-10 |
| 4 | 1-3 | 9-11 |
| 5 | 0-2 | 9-11 |
| 6 | 0-2 | 9-11 |
| 7 | 0-2 | 9-11 |
| 8 | 1-3 | 9-11 |
| 9 | 0-2 | 8-10 |
| 10 | 0-2 | 8-10 |
| 11 | 0-2 | 8-10 |
| 12 | 0-2 | 8-10 |

**Table S1.** Recording contacts for each STN for all participants
