## Supplementary Information, Fig. S1-8 for "Differential Effects of Pathological Beta Burst Dynamics Between Parkinson’s Disease Phenotypes Across Different Movements"

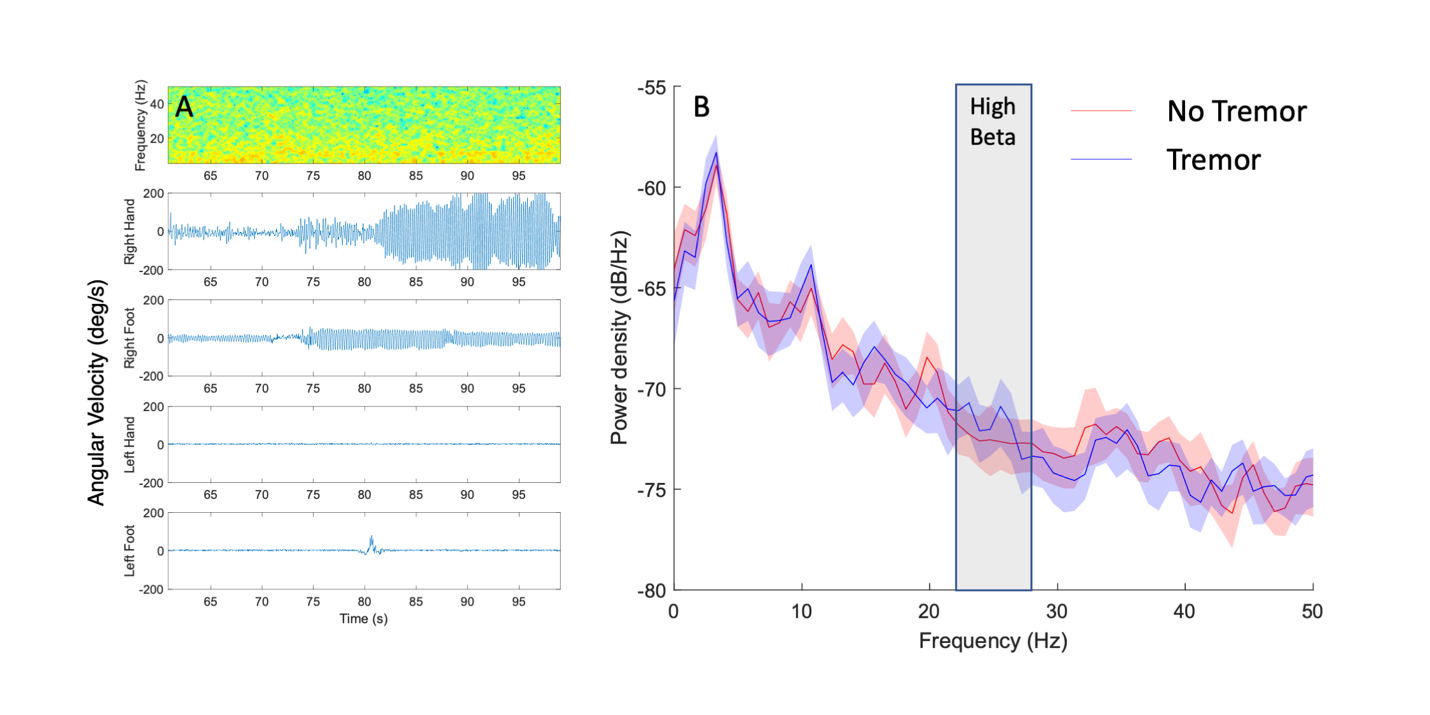

**Figure S1.** Example figure of Participant 3 (tremor-dominant) during the QDG task (right hand, left STN). **(A)** Synchronized spectrogram of neural data and movement of each limbed measured by a gyroscope. **(B)** Power spectral density plots of periods of no tremor and tremor in the right hand.

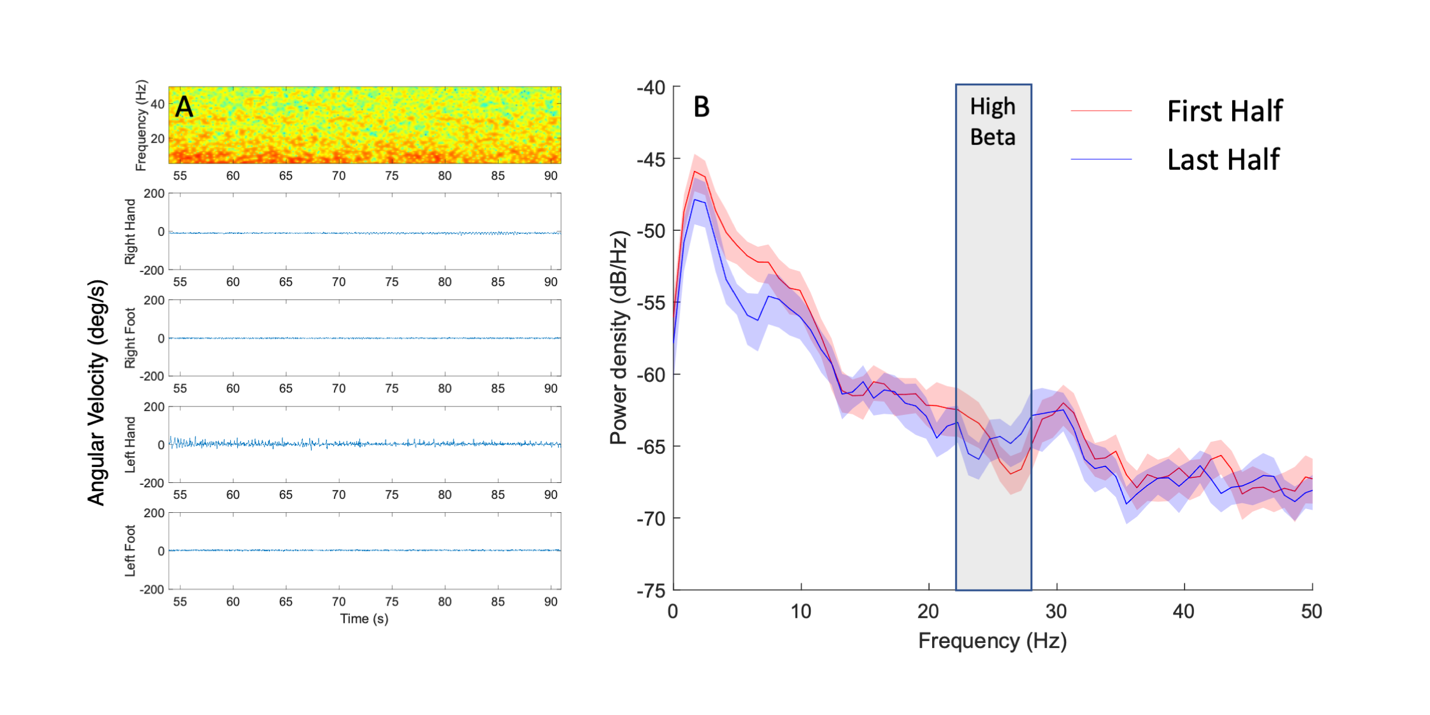

**Figure S2.** Example figure of Participant 3 (tremor-dominant) during the QDG task (left hand, right STN). **(A)** Synchronized spectrogram of neural data and movement of each limbed measured by a gyroscope. **(B)** Power spectral density plots of the first and last half of trial as there was no onset of tremor in the middle of the trial for the left body.

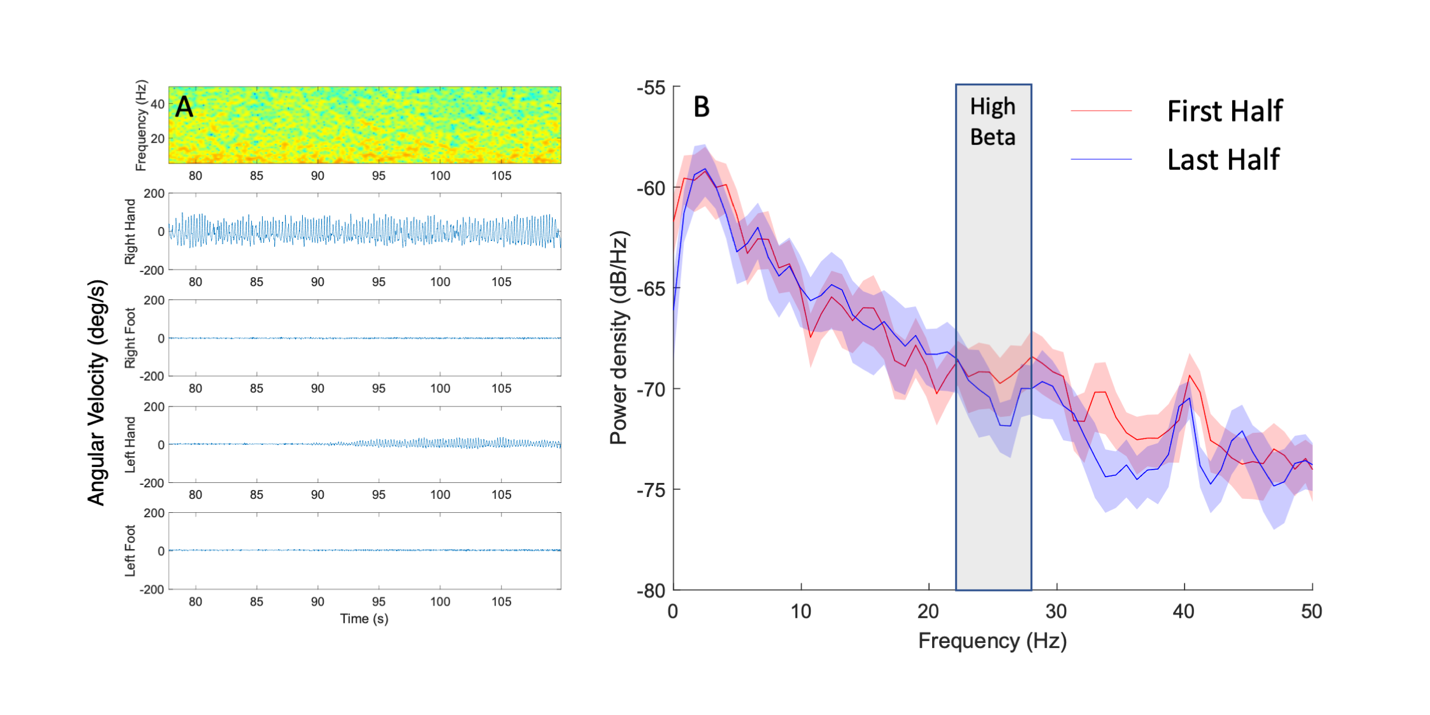

**Figure S3.** Example figure of Participant 6 (tremor-dominant) during the QDG task (right hand, left STN). **(A)** Synchronized spectrogram of neural data and movement of each limbed measured by a gyroscope. **(B)** Power spectral density plots of the first and last half of trial as there was no onset of tremor in the middle of the trial for the right body.

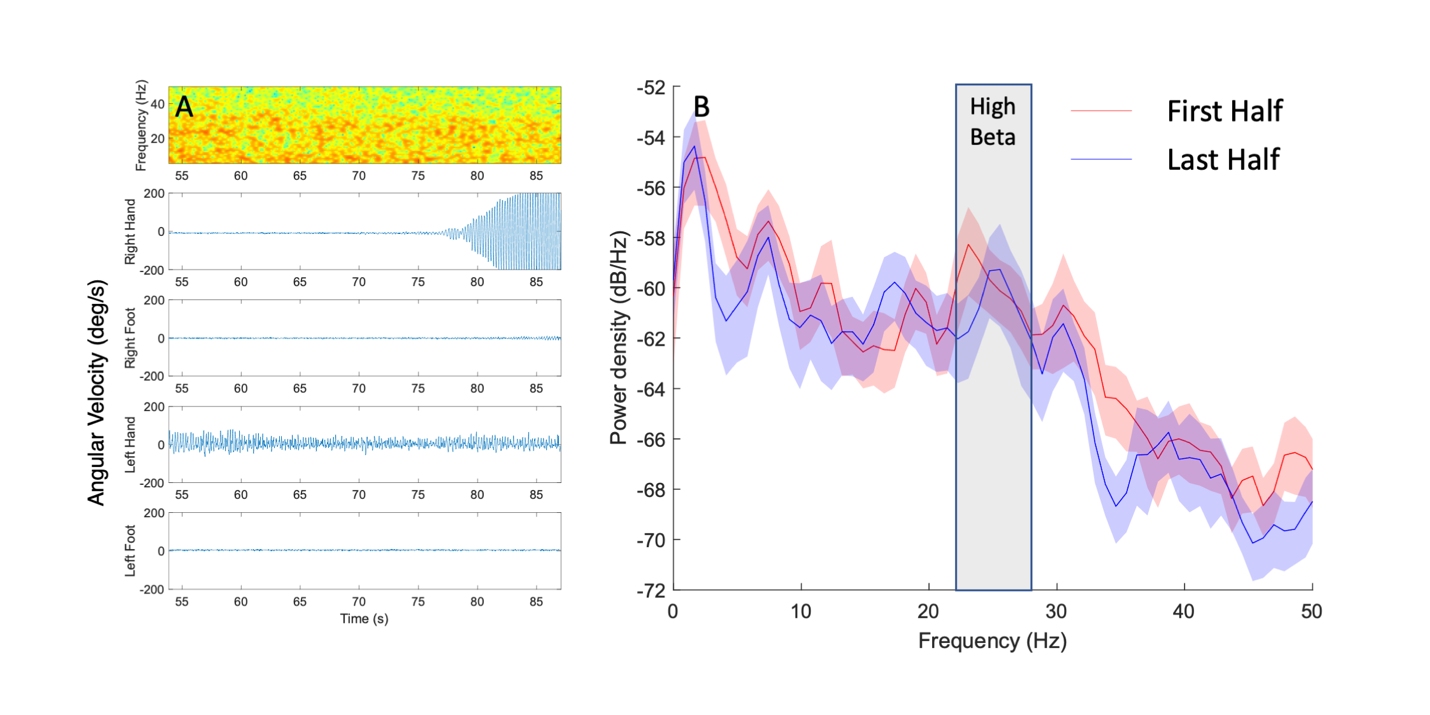

**Figure S4.** Example figure of Participant 6 (tremor-dominant) during the QDG task (left hand, right STN). **(A)** Synchronized spectrogram of neural data and movement of each limbed measured by a gyroscope. **(B)** Power spectral density plots of the first and last half of trial as there was no onset of tremor in the middle of the trial for the left body.

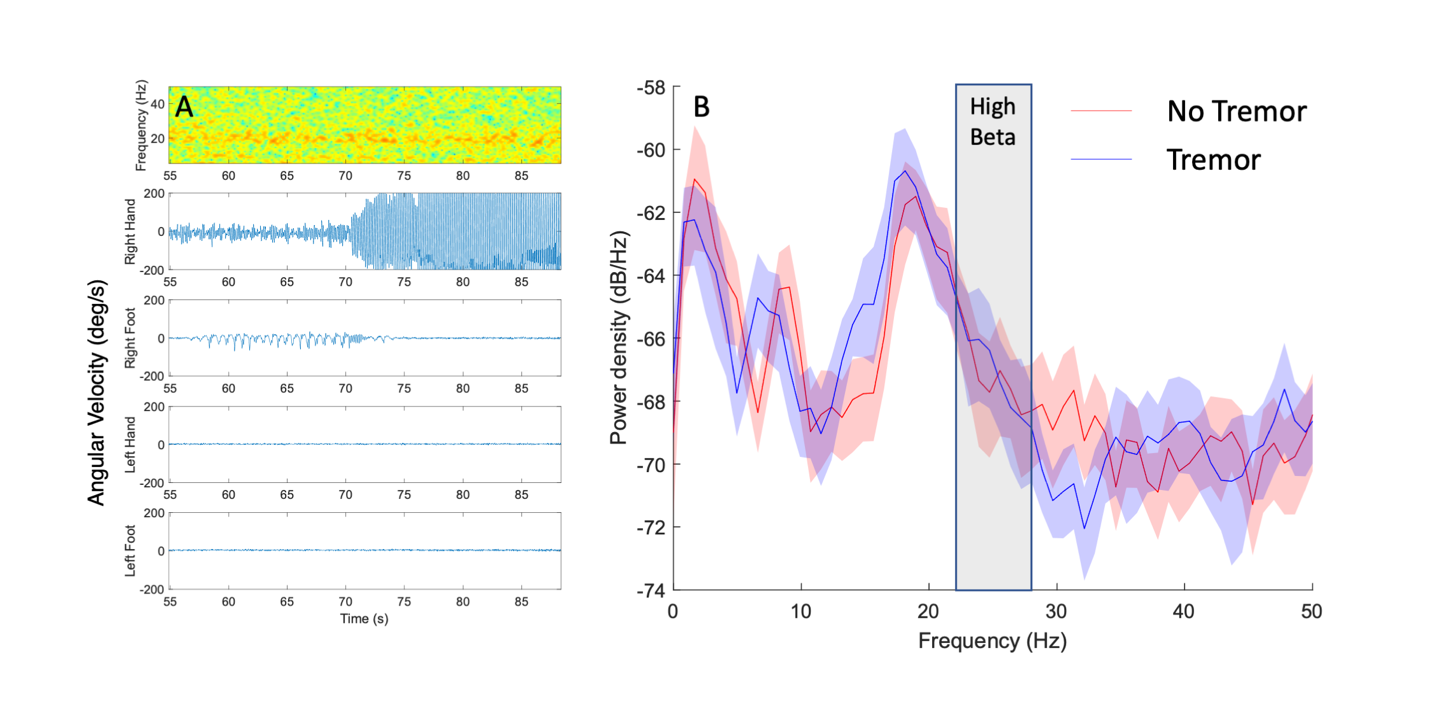

**Figure S5.** Example figure of Participant 8 (tremor-dominant) during the QDG task (right hand, left STN). **(A)** Synchronized spectrogram of neural data and movement of each limbed measured by a gyroscope. **(B)** Power spectral density plots of periods of no tremor and tremor in the right hand.

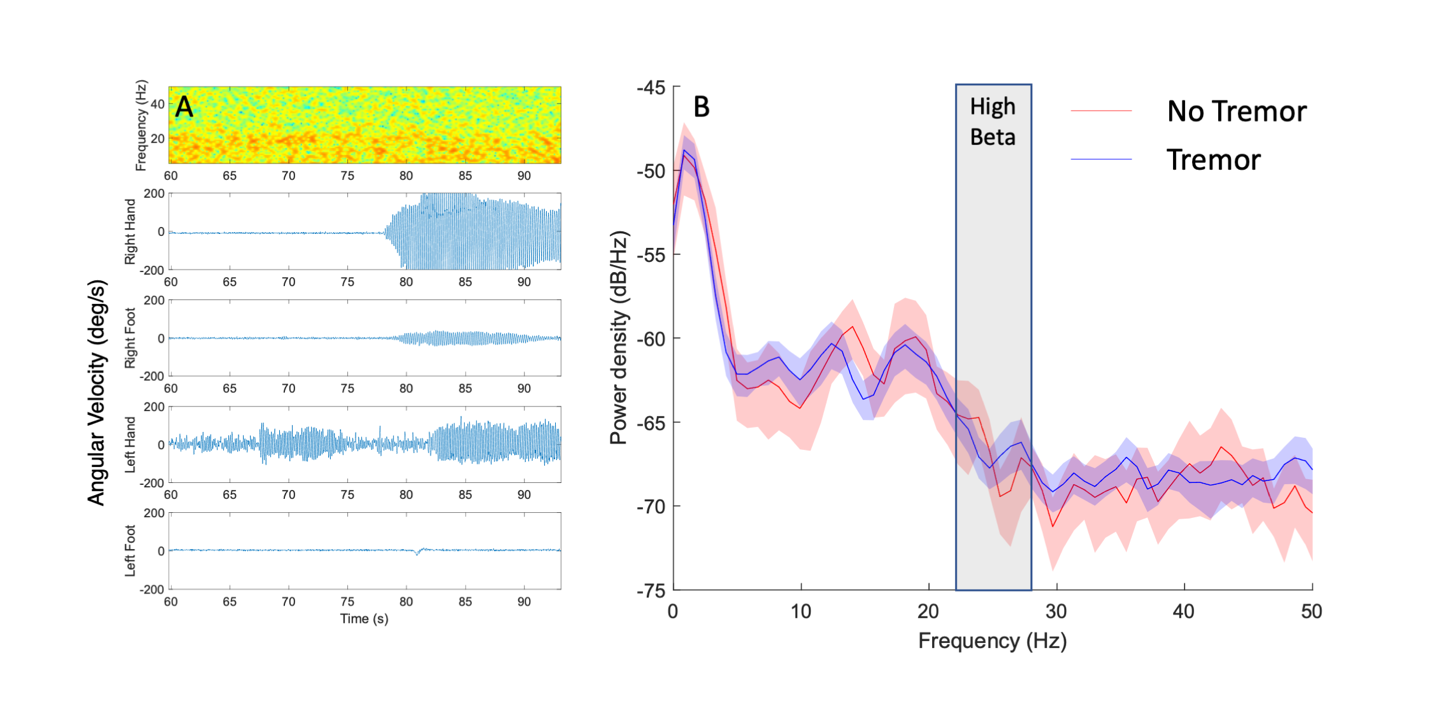

**Figure S6.** Example figure of Participant 8 (tremor-dominant) during the QDG task (left hand, right STN). **(A)** Synchronized spectrogram of neural data and movement of each limbed measured by a gyroscope. **(B)** Power spectral density plots of periods of no tremor and tremor in the left hand.

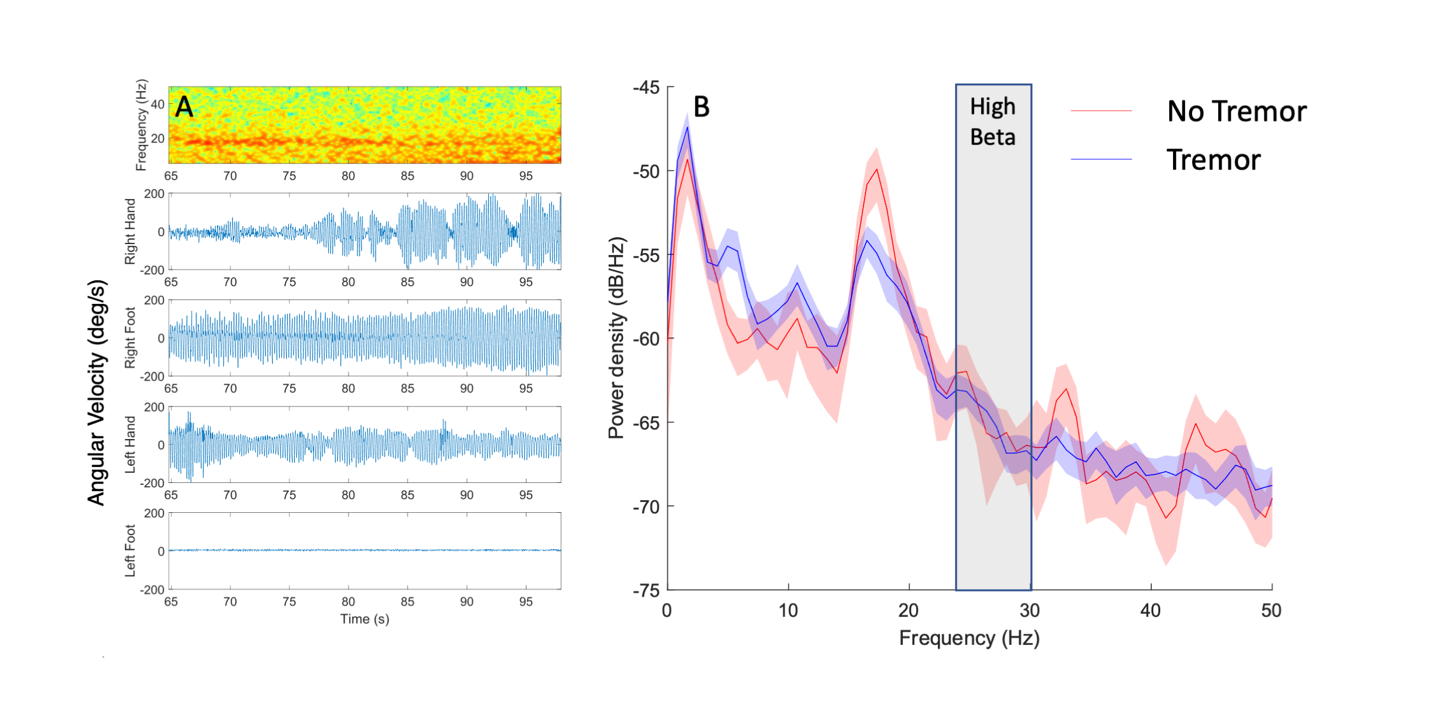

**Figure S7.** Example figure of Participant 12 (tremor-dominant) during the QDG task (right hand, left STN). **(A)** Synchronized spectrogram of neural data and movement of each limbed measured by a gyroscope. **(B)** Power spectral density plots of periods of no tremor and tremor in the right hand.

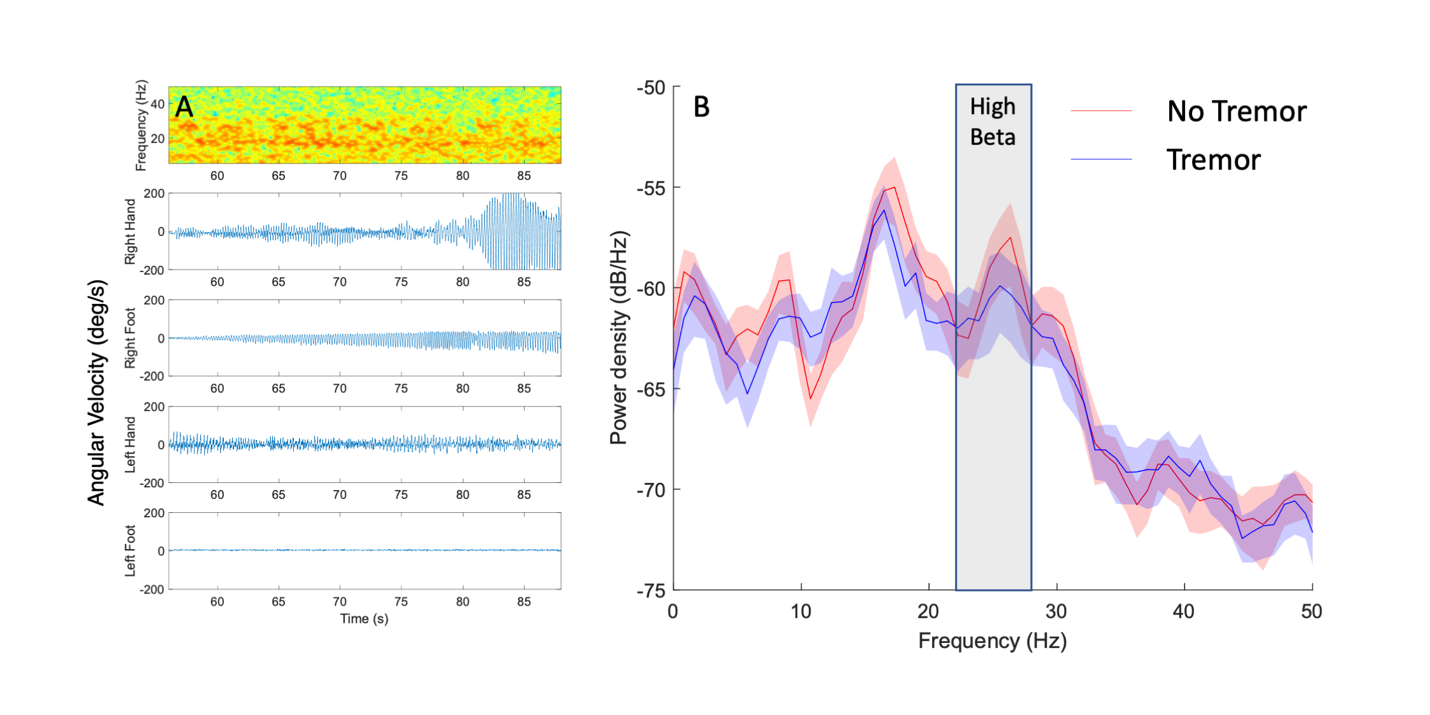

**Figure S8.** Example figure of Participant 12 (tremor-dominant) during the QDG task (left hand, right STN). **(A)** Synchronized spectrogram of neural data and movement of each limbed measured by a gyroscope. **(B)** Power spectral density plots of periods of no tremor and tremor in the left hand.
